# Immune Cell-Based Transcriptomic Mendelian Randomization and Colocalization Study on Type 1 Diabetes

**DOI:** 10.1101/2025.07.01.25330014

**Authors:** Julie Sklar, David Stacey, Grace Nickel, Adam S Butterworth, Elias Allara, Liam Gaziano

## Abstract

**Background:** Interventions to prevent type 1 diabetes (T1D), a disease requiring lifelong treatment, remain limited. We sought to identify novel therapeutic targets for T1D through Mendelian randomization and colocalization using immune cell-derived instruments.

**Methods:** We selected locally-acting genetic variants from 14 transcriptomic studies to instrument expression of 8,998 genes measured in immune cell types. Outcome associations were obtained from a T1D genome-wide association study that included 18,942 cases and 501,638 controls. Follow-up analyses included assessments of horizontal pleiotropy and novelty, as well as phenome-wide scans with colocalization (PheWAS-coloc) of instrumental variants.

**Results:** We prioritized 21 genes (*CLNK*, *EED*, *LZTFL1*, *MGAT4A*, *NAA38*, *NFKB1*, *PHACTR4*, *PHLPP2*, *PLEKHA1*, *P2RY12*, *REST*, *RGS14*, *SERPINB6*, *SESN3*, *SLC25A29*, *SPAG1*, *STIM2*, *THEMIS*, *TMEM80*, *VSIR*, *ZNF217*) that have not been identified in previous T1D genome-wide association studies. Notably, higher genetically-predicted *VSIR* (encoding the immune checkpoint protein VISTA) was associated with decreased risk of T1D, providing human genetic support that complements existing animal-model evidence for VISTA’s protective role in autoimmune diseases. We also prioritized *P2RY12*, which is targeted by several approved drugs, revealing a possible repurposing opportunity. PheWAS-coloc analyses further linked *P2RY12* expression to Epstein-Barr virus EBNA-1 antibody levels, implicating a pathway relevant to autoimmunity.

**Conclusions:** Our findings provide novel avenues for drug development and repurposing to prevent or delay T1D onset.

## Introduction

Type 1 diabetes (T1D) is caused by immune-mediated loss of insulin-producing β cells in pancreatic islets. Often diagnosed in childhood, T1D requires lifelong, daily management and is associated with premature morbidity and mortality^1^. Beyond its clinical impact, T1D imposes considerable societal costs, including direct medical expenditures and lost productivity^2^, which are likely to increase as the incidence of T1D rises^3^. Therefore, preventing disease onset would greatly reduce the financial and health burdens associated with T1D on the individual and societal level.

The trial of teplizumab (an anti-CD3 antibody) showed that targeted therapy against a single protein can delay disease onset in high-risk relatives of patients with T1D^4^. There is a need for additional modalities beyond teplizumab, though, since 43% of those receiving the drug still developed T1D over a median follow-up of two years. Mendelian randomization (MR) techniques can help identify potential therapeutic targets by assessing their causal role in disease^5^. MR leverages the randomization scheme introduced during segregation of alleles to help avoid influences from confounding and reverse causation that are inherent in observational data^6^. Moreover, findings from human genetic data may be more applicable to human biology than functional work in animal models. Indeed, therapeutics targeting proteins with supportive human genetic evidence are more likely to gain market approval^7^.

Previous MR studies on T1D have provided insights into etiology of T1D by leveraging instrumental variants that influence levels of plasma proteins^8,9^. While informative, blood plasma may not be as relevant to the pathogenesis of T1D compared to other tissues or cell types, like immune cells. Therefore, to identify potential new drug targets for T1D, we performed a transcriptomic-based MR study on T1D using gene expression quantitative trait locus (eQTL) instruments in isolated immune cells.

## Methods

### Mendelian Randomization Assumptions

MR relies on three key assumptions: relevance, independence, and exclusion restriction. To satisfy the relevance assumption, instrumental variants should be robustly associated with the exposure of interest. For the independence assumption, instrumental variants should not be associated with any confounders. For example, without population stratification a variant may have higher allele frequency in one ancestry, and that ancestry could be enriched for disease, confounding the association between the variant and disease. For exclusion restriction, instrumental variants must exert their effect through the instrumented exposure, and not through horizontally pleiotropic pathways.

### Genetic instruments and outcome associations

To select genetic instruments, we used 29 eQTL datasets from 14 studies^10-23^ that measured gene expression in immune cell types (full information for each study can be found in **Supplementary Table 1**), downloaded from the eQTL Catalogue^24^, a resource that aggregates publicly available data on gene expression QTLs. We extracted all locally-acting (*cis*) eQTLs with *P*<5×10^-8^ (to meet the relevance assumption), and conducted LD clumping (r^2^<0.1) using plink (v1.07) to select approximately independent *cis* instruments for each gene within each dataset. We restricted the analysis to protein coding genes (i.e. those with a UniProt identifier) and removed variants that lie within the major histocompatibility complex (MHC) region (chr6:25,726,063-33,400,644), due to the complex genetic correlation structure in that region^8,25^. Associations between instrumental variants and T1D were obtained from a GWAS meta-analysis described in Chiou *et al*.^26^, which included 18,942 T1D cases and 501,638 controls of European ancestry and is the largest publicly available GWAS to date. Though this GWAS is limited by its restriction to European individuals, it helps fulfill the independence assumption of MR because eQTL instruments were derived in primarily European populations as well.

### Mendelian Randomization Analysis

We used the Wald ratio method to perform MR for instruments with one variant. For instruments with more than one variant (i.e., two independent variants are associated with the same gene in the same dataset), we performed fixed-effect inverse variance-weighted MR, which involves meta-analyzing variant-level MR effects. We also evaluated heterogeneity across the variant-level MR estimates and presented the Cochran Q P-value for instruments with multiple variants. A Bonferroni-adjusted significance threshold corrected for the number of genes within each of the 29 datasets was used (**Supplementary Table 1)**. Analyses were performed using the TwoSampleMR R package https://github.com/MRCIEU/TwoSampleMR)

### Pairwise Conditional Colocalization

Significant MR results can arise because an instrumental variant is correlated (in linkage disequilibrium [LD]) with a causal variant in a region, but the instrumental variant is not causally influencing disease itself, also known as confounding by LD. To assess the possibility this is occurring, we performed conditional colocalization (PWCoCo)^27^ between the eQTL summary statistics (±500kb around instrumental variants) and T1D for all results that passed the MR P-value thresholds. Colocalization uses a Bayesian approach to calculate the five posterior probabilities: Posterior Probability of Hypothesis 0 (PPH0) is the probability that a signal does not exist for either gene expression or T1D; PPH1, a signal exists for gene expression, but no signal exists for T1D; PPH2, a signal exists for T1D, but no signal exists for gene expression; PPH3, a signal exists for gene expression and T1D, but the two signals are independent of each other; PPH4 a shared signal exists for both gene expression and T1D. We used the PWCoCo default priors (p1=1×10^-4^, p2=1×10^-4^, p12=5×10^-5^) and a colocalization threshold of PPH4 > 0.8. PWCoCo initially performs colocalization using marginal summary statistics (marginal colocalization), then integrates the GCTA-COJO^28^ program to perform conditional analysis using LD information (UK Biobank was used as a reference panel in the present analysis), if the PPH4 is less than 0.8 using marginal colocalization. This conditioning step can remove strong, independent signals that interfere with colocalization, as it assumes a single causal signal per region tested.

### Follow-up analyses

#### Assessment of horizontal pleiotropy

Restricting analyses to *cis*-eQTLs increases the likelihood that associations between eQTLs and phenotypes represent downstream consequences (vertical pleiotropy), which do not violate the exclusion restriction assumption. However, an eQTL may lie in a regulatory region that influences expression of other nearby genes. In such a case, it can be difficult to determine which of the genes associated with the instrumental variant is truly driving the association with the outcome, a phenomenon that we consider a form of horizontal pleiotropy. For example, in the 1q23.1 region, eQTLs for both *TTC24* (in regulatory-T-Cell memory cells) and *GPATCH4* (in CD4+ T cells) colocalize with the same T1D signal, and rs35091159 is an instrumental variant for both genes. Therefore, we grouped results that passed MR and colocalization thresholds by region, defined as when instrumental variants from different genes were in close proximity (+250kb). Results in regions positive for multiple unique genes were considered to exhibit horizontal pleiotropy.

#### Assessment of novelty

We classified results as novel if they did not include instrumental variants that were correlated (*r*^2^<0.2 in 1000 Genomes EUR) or in close proximity (+250kb) with genome-wide significant (P<5×10^-8^) variants for T1D reported in five previous association studies^26,29-32^, and not identified in two previous MR studies^8,9^.

#### Phenome-wide scans with Colocalization

PheWAS can reveal on-target effects on outcomes beyond T1D, like safety concerns, that we would want to anticipate, and it can uncover possible meditating phenotypes biologically relevant to T1D development, that can reveal mechanistic insights and add to the credibility of a result. Therefore, for results that were novel and the lone result regionally, we queried instrumental variants in the MRbase PheWAS platform^33^, a database that contains over 50,000 publicly-available GWAS on numerous traits and outcomes. We also queried variants for association with plasma levels of 4,775 proteins measured by the SomaLogic platform within 10,708 individuals^34,35^ (data available at www.omicscience.org). For genes with instruments that contain multiple independent variants, we selected the variant most strongly associated with gene expression for PheWAS. We displayed associations with *P*<5×10^-5^ and also employed PWCoCo between eQTLs and PheWAS summary stats ±500kb around the queried variant, to help assess whether PheWAS associations are due to confounding by LD. It is worth noting that these PheWAS-coloc analyses can be an additional tool to uncover shared causal associations between instrumental variants and expression of nearby genes or proteins (horizontal pleiotropy), through the SomaLogic protein datasets queried and the MR-base platform, which includes GWAS on whole blood expression of nearly 20,000 genes from the eQTLGen Consortium^36^.

## Results

### Study design

**Figure 1** shows the overall study design. We selected locally-acting eQTLs (*cis*-eQTLs) for gene expression in immune-related cells from 14 studies (29 datasets) within the eQTL catalogue^24^. More information on each study is provided in **Supplementary Table 1**. Outcome data came from a GWAS on T1D that included 18,942 cases and 501,638 controls^26^. For results that passed MR significance (P<0.05 Bonferroni-corrected for unique number of genes tested within each of the 29 datasets), we additionally performed Pairwise Conditional Colocalization (PWCoCo)^27^ to assess for confounding by LD.

**Figure 1.**
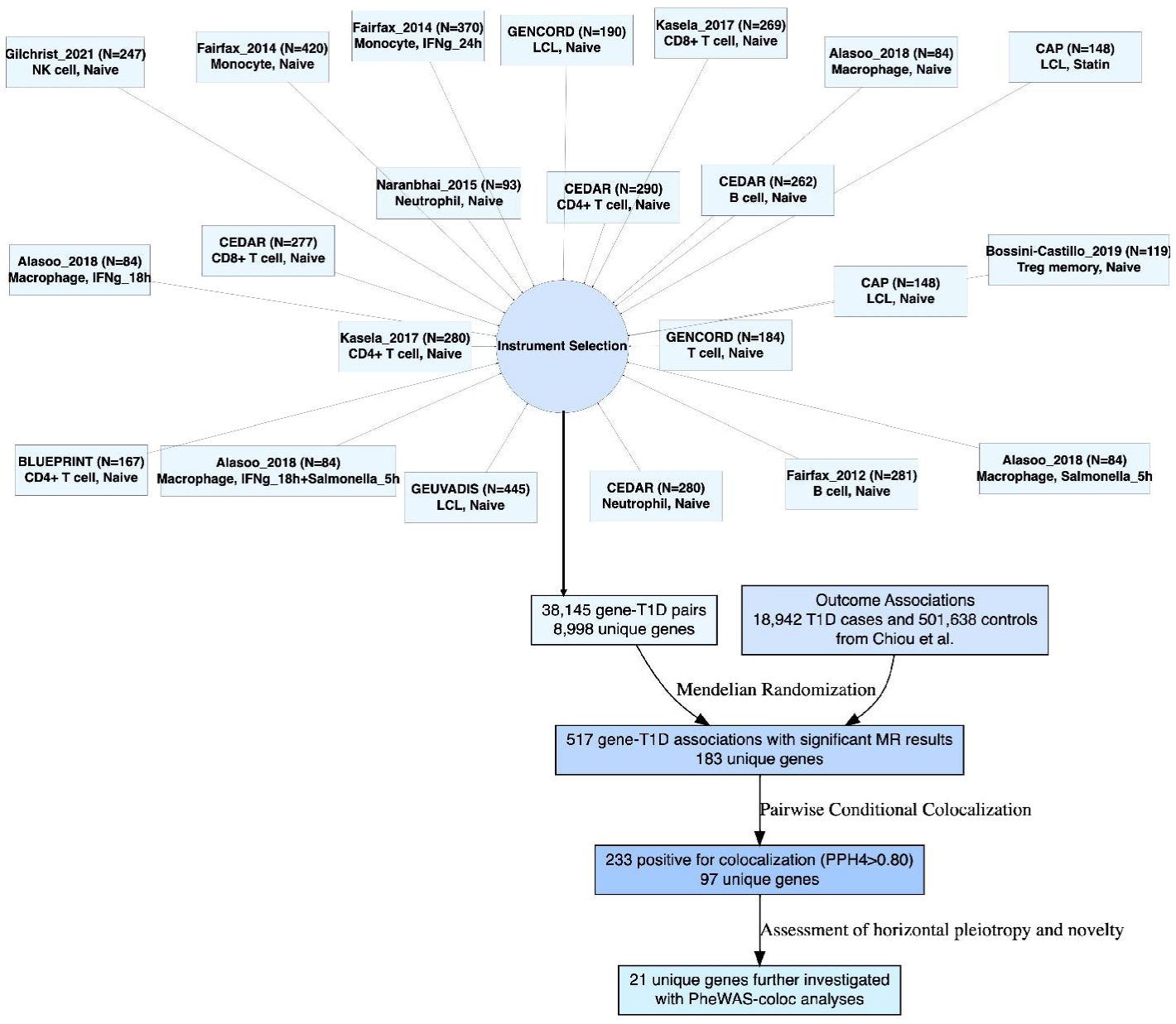
Overall study design. Displayed are the 29 datasets (with study, sample size, immune cell type and cell conditions) that were used to select locally-acting eQTLs (*cis*-eQTLs) for gene expression as instrumental variants. Associations with T1D were obtained from Chiou et *al*.^26^. Conditional colocalization was performed with PWCoCo^27^. PheWAS-coloc refers to the phenome-wide scans with colocalization described in Section 2.4.

### Mendelian randomization and colocalization

Across 29 datasets, 38,145 gene-T1D associations were tested. 90.6% of instruments included one *cis*-eQTL, 4.1% had two, 4.3% had three, 1.0% had four or more. Two genes had instruments available in all 29 eQTL datasets, while 2,478 genes had instruments available in only one dataset (**Supplementary Table 2**). 517 gene-T1D associations passed MR significance (**Supplementary Table 3**), 233 of which also passed colocalization (Figure 2**, Supplementary Table 4**). In terms of unique genes, 8,998 were tested, 183 passed MR significance and 97 genes additionally colocalized (**Figure 2, Supplementary Table 5**).

**Figure 2.**
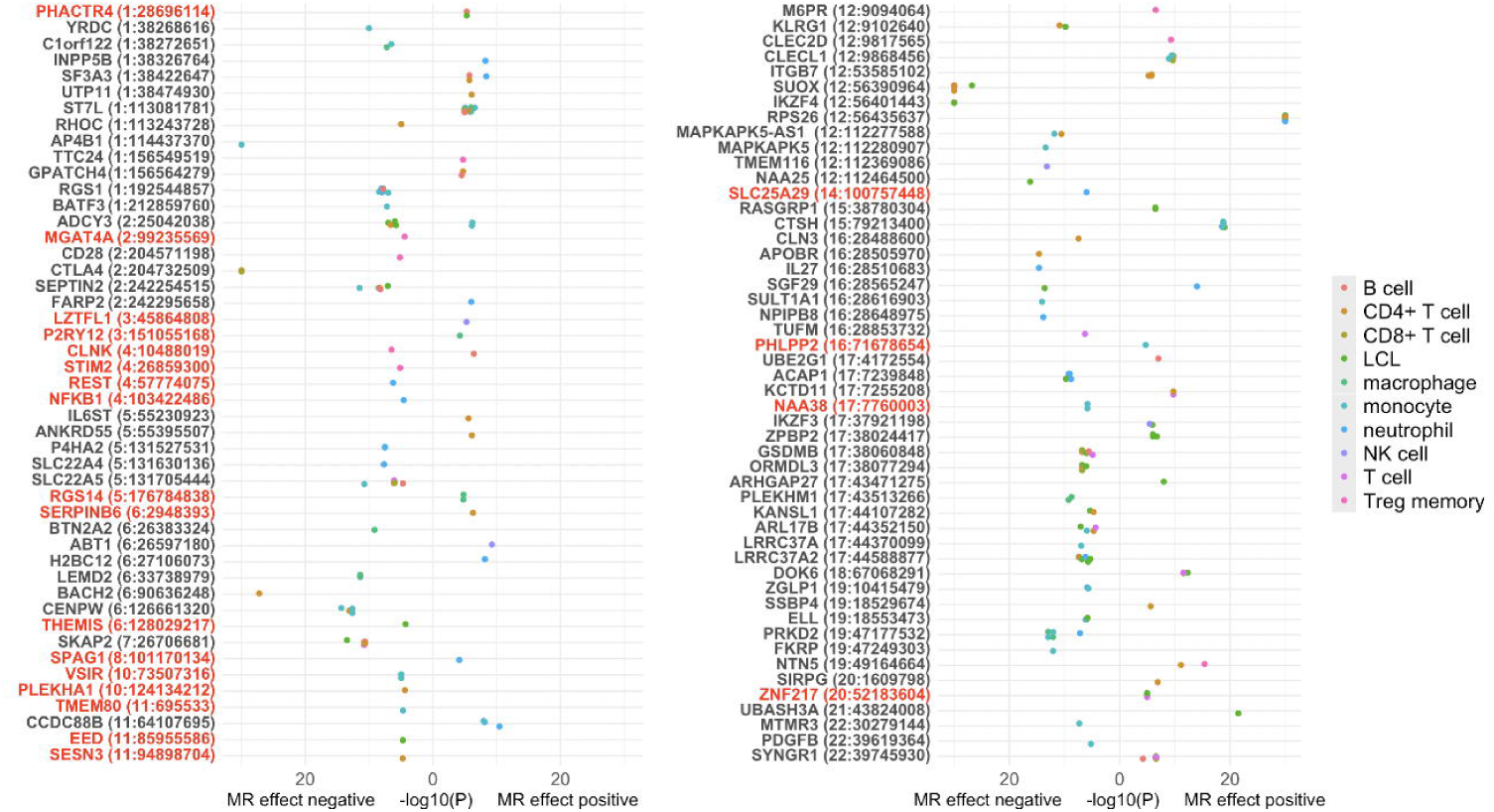
Bidirectional plot of results that passed both Mendelian randomization and colocalization thresholds. Shown are associations for 233 gene-T1D pairs grouped by unique gene (chromosome: transcription start site), with color according to immune cell types used to derive instrumental variants. -log_10_(P-value) from MR analysis is plotted to the right if the MR effect-estimate was positive and to the left if negative. -log_10_(P-value) is truncated at 30. Genes in red indicate ones that are both the lone result in the region and novel. LCL, lymphoblastoid cell line. NK, natural killer. Treg memory, regulatory-T-Cell memory cells.

There was possible enrichment for results using instruments derived in B and T cells compared with other immune cell types. There were 1,757 unique genes tested in B-cells of which 17 (0.97%) passed MR thresholds and colocalization thresholds, 0.95% passed for T cells, 0.77% for regulatory-T-Cell memory cells (Treg memory), 0.75% for neutrophils, 0.72% for lymphoblastoid cell line (LCL), 0.67% for macrophages, 0.61% for monocytes, 0.36% for natural killer (NK) cells, and 0% for microglia. The highest percentage of unique genes that passed colocalization of those that passed MR significance was in B cells (56.7%) and T cells (54.9%), and the lowest was in NK cells (28.6%) and microglia (0%, **Figure 3**).

**Figure 3.**
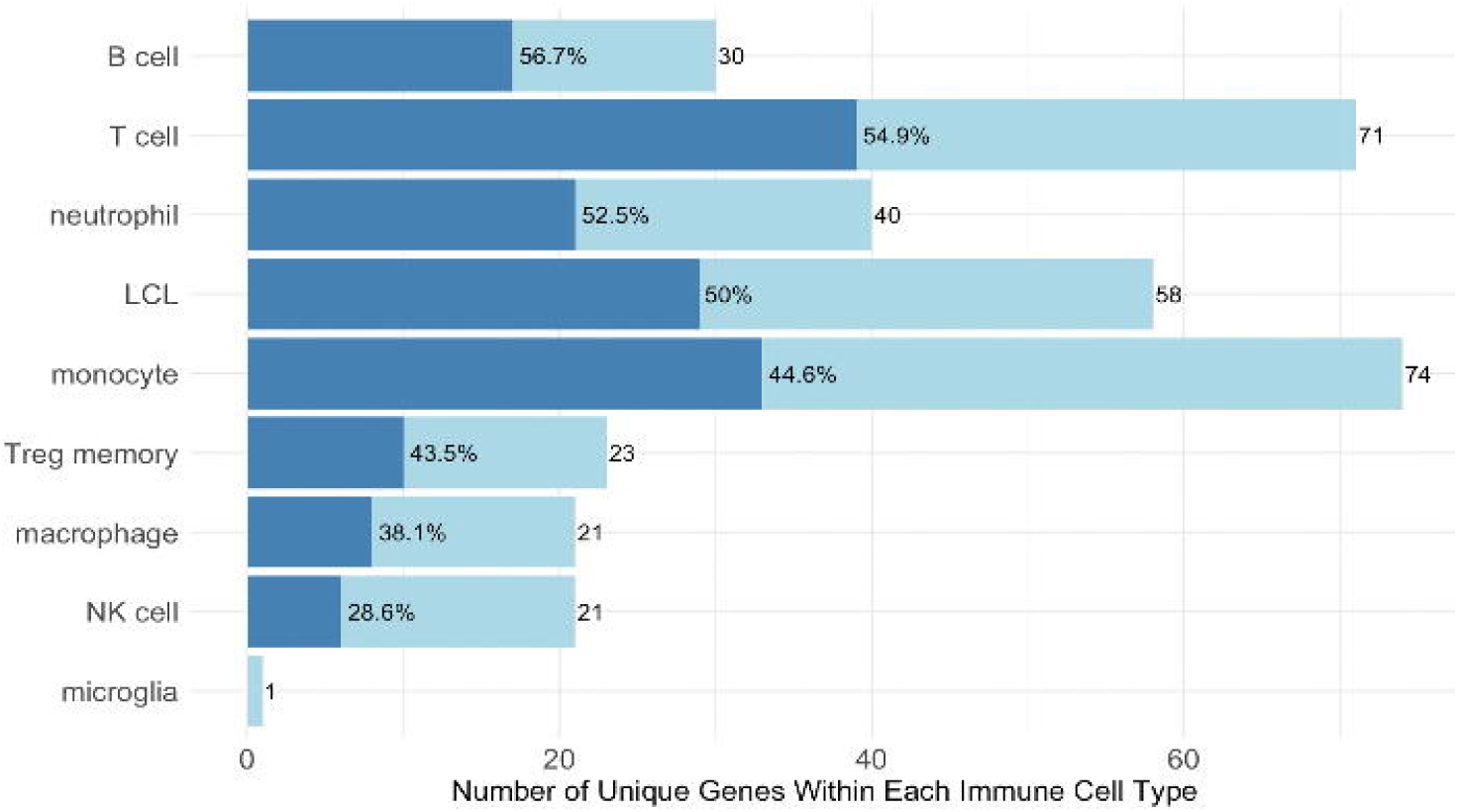
Genes passing Mendelian randomization significance and colocalization by immune cell type. Light blue bars represent the number of unique genes that are significant in each cell type, and dark blue bars represent the number of unique genes that additionally passed colocalization. Results from eQTL datasets derived in the same immune cell type (CD4+ T Cells, CD8+ T Cells, and T Cells were considered one cell type) were combined before calculating unique number of genes. LCL, lymphoblastoid cell line. NK, natural killer. Treg memory, regulatory-T-Cell memory cells.

### Prioritizing genes that are novel and the lone result in a region

We considered 27 of the 97 colocalizing genes as novel, as they have not been identified in previous T1D GWAS or MR studies (**Supplementary Table 5**). Of the 97 colocalizing genes, 56 were located in close proximity (±250kb) to at least one other result, and 41 were the sole result in a region (**Supplementary Table 5**). Twenty-one genes (*CLNK*, *EED*, *LZTFL1*, *MGAT4A*, *NAA38*, *NFKB1*, *PHACTR4*, *PHLPP2*, *PLEKHA1*, *P2RY12*, *REST*, *RGS14*, *SERPINB6*, *SESN3*, *SLC25A29*, *SPAG1*, *STIM2*, *THEMIS*, *TMEM80*, *VSIR*, *ZNF217*, **Figure 2**) were both the lone result in the region and novel. There were instruments for *ZNF217* in six eQTL datasets, all of which were significantly associated with T1D through MR, and five of which additionally colocalized (unique cell types: B cell, LCL, T cell, monocyte, **Figure 4**). By contrast, 16 *TMEM80*-T1D associations were tested, only one of which passed MR and colocalization thresholds.

**Figure 4.**
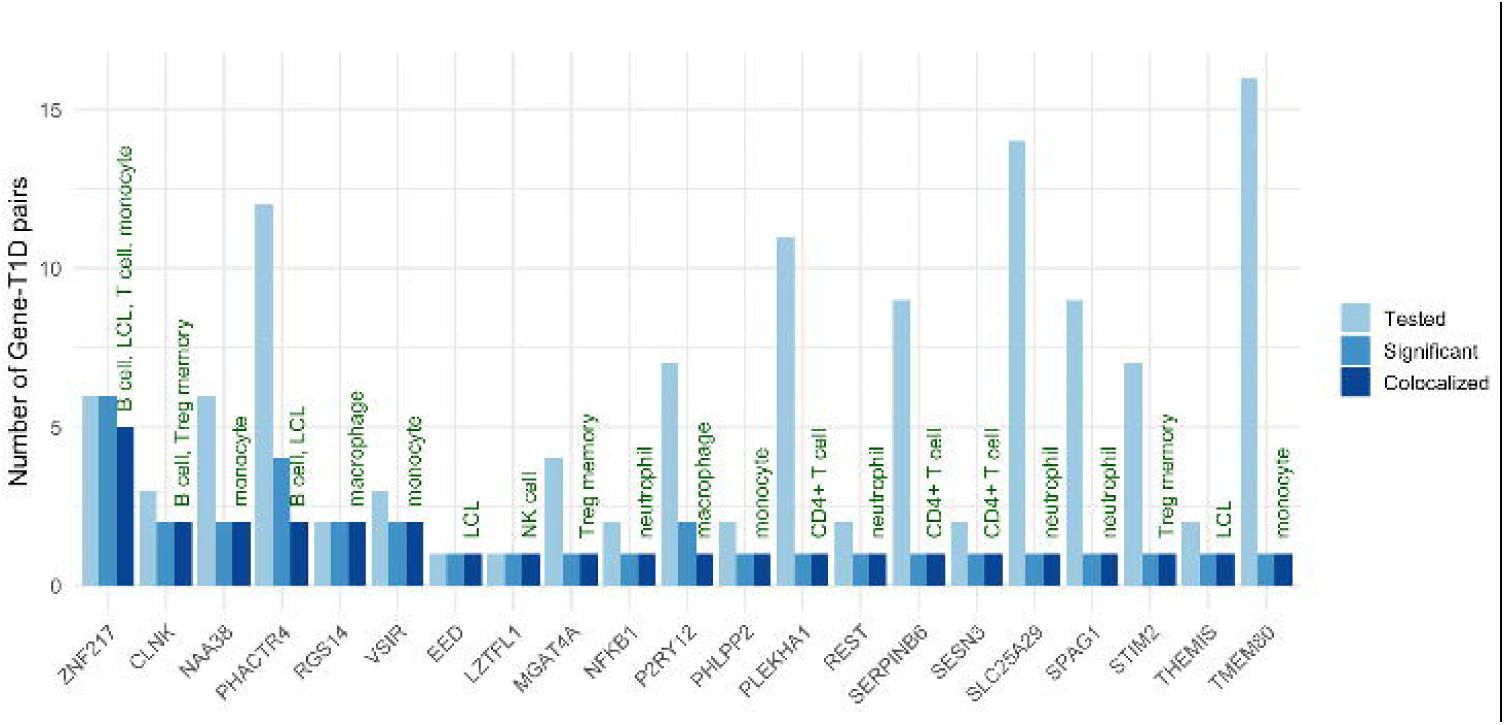
Prioritized genes across eQTL datasets. Light blue bars indicate the number of gene-T1D MR associations that were tested (i.e. the number of datasets in which a *cis*-eQTL instrument existed), medium blue bars indicate the number that were MR significant, and dark blue indicates the number that additionally colocalized. Green labels indicate the unique immune cell types used to derive eQTL instruments that passed MR and colocalization thresholds. LCL, lymphoblastoid cell line. NK, natural killer. Treg memory, regulatory-T-Cell memory cells.

### Phenome-wide scans of instrumental variants with conditional colocalization

We then performed phenome-wide scans of instrumental variants with conditional colocalization (PheWAS-coloc) for the 21 prioritized genes. Several genes had PheWAS-coloc results (associated at P<5×10^-5^ and colocalized at PPH4 > 0.8) for immune-mediated conditions beyond T1D, including primary sclerosing cholangitis (*CLNK* and *NFKB1*), myasthenia gravis (*SESN3*), Grave’s disease (*CLNK* and *SESN3*), rheumatoid arthritis (*CLNK* and *SESN3*), and asthma (*EED*, *NFKB1*, *REST*). Full PheWAS-coloc associations for the 21 prioritized genes can be found in **Supplementary Table 6**.

Select PheWAS-coloc results for *VSIR* (eQTLs derived in monocyte cells) and *P2RY12* (macrophages), *CLNK* (Treg memory) and *SESN3* (T cells) are shown in **Figure 5**, where MR-estimates correspond to higher genetically predicted gene expression. Genetically-predicted *VSIR* was associated with plasma levels of C-X-C motif chemokine 16 (CXCL16, encoded by *CXCL16* on chromosome 17) and C-C motif chemokine 17 (CCL17, encoded by *CCL17* gene on chromosome 16). Because these proteins represent trans associations, encoded by genes distantly located from *VSIR*, they could represent consequences of *VSIR* function, as opposed to direct influences of the *cis*-eQTL on protein expression (i.e., more likely vertical pleiotropy rather than horizontal). *VSIR* was also positively associated with basal cell carcinoma (BCC) and inversely associated with stool abundance of a strain of Blautia, an anaerobic bacteria found in the intestine of animals^37^. PheWAS-coloc for *P2RY12* showed a positive association with Epstein-Barr virus EBNA-1 antibody levels. It also revealed the possibility of horizontal pleiotropy through shared expression of nearby genes, in that eQTLs for *P2RY12* colocalized with eQTLs for the nearby gene *SUCNR1*. However, eQTLs for *P2RY12* only colocalized with *SUCNR1* after conditional analysis, an indication that *P2RY12* is prioritized in this region. *SESN3* and *CLNK* were both associated multiple autoimmune diseases. Higher genetically-predicted *SESN3* was additionally associated with lower levels of CD3 on resting CD4 regulatory T cells and CD28 on CD45RA+ CD4+ T cells.

**Figure 5.**
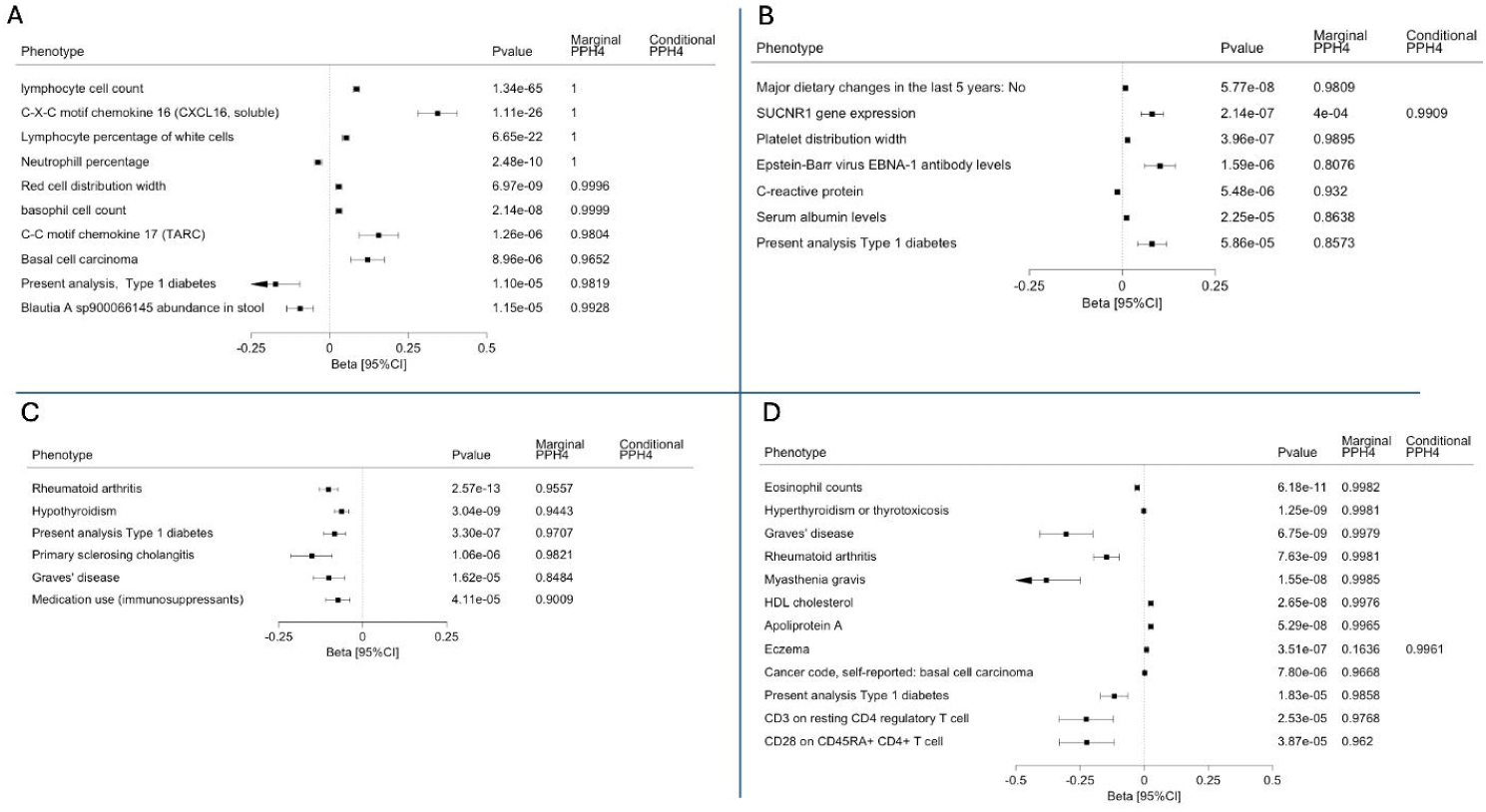
PheWAS-coloc results for (A) *VSIR*, (B) *P2RY12*, (C) *CLNK, and (D) SESN3*. PheWAS associations for **(A)** rs748113, the *cis*-eQTL for *VSIR* in monocyte cells, **(B)** rs10513398 the *cis*-eQTL for *P2RY12* in macrophage cells **(C)** rs9291444 the *cis*-eQTL for *CLNK* in regulatory-T-Cell memory cells, and **(D)** rs4409785, the *cis*-eQTL for *SESN3* in T cells. Beta estimates are variant-level MR estimates for higher genetically predicted gene expression. Instrumental variants were queried for traits/outcomes in the MRBase PheWAS platform, and plasma proteins reported in omicscience.org. Colocalization was performed ±500kb around instrumental variants between eQTLs and PheWAS summary statistics using pairwise conditional colocalization (PWCoCo). Shown are associations with P<5×10^-5^ and colocalization PPH4>0.80. Full PheWAS-coloc results can be found in **Supplementary Table 6**. Marginal PPH4, posterior probability of hypothesis 4 using marginal summary statistics. Conditional PPH4, colocalization with conditional analysis.

## Discussion

Preventing T1D could spare millions of individuals from its negative impacts on quality of life, morbidity and mortality. To identify potential therapeutic targets, we leveraged genetic instruments derived in immune cells to perform MR and colocalization for nearly 9000 genes on T1D. We prioritized 21 genes that have not been identified in previous GWA or MR studies, and uncovered important insights through PheWAS-coloc analyses. Collectively, these findings highlight new avenues for development or repurposing of drugs to prevent or delay the onset of T1D.

An intriguing result is *VSIR*, which encodes the V-domain immunoglobulin suppressor of T cell activation protein (known as GI24 or VISTA), because of its biological role in immune cell regulation. VISTA is an immune checkpoint protein expressed on myeloid cells that limits immune response against self-antigen^38^. It is a member of the B7 family ligands, like programmed death-ligand 1 (PD-L1), that reduces the CD4+ and CD8+ T cell activation^39^. A growing body of evidence exists on the protective role of VISTA in autoimmune disease, primarily through mouse models. VISTA knockout mice (VISTA-/-) had an increased incidence and intensity of experimental autoimmune encephalomyelitis, systemic lupus erythematosus (SLE), and inflammatory lupus when bred with mice that are predisposed to developing autoimmune disease^40^. Administration of an anti-VISTA antibody resulted in accelerated disease severity in a mouse model for multiple sclerosis^41^. Conversely, agonism of VISTA in mouse models of SLE^42^ and psoriasis^43^ improved disease. Here, we demonstrate that the effect of *VSIR* on autoimmunity, previously shown in animal models, is corroborated by human genetic data – an important predictor for clinical success of a targeting therapeutic. Additionally, modulation of VISTA to prevent or delay T1D is feasible in the near-term, as an oral small molecule VISTA agonist (M351-0056) has been identified^43^.

The PheWAS-coloc analysis provided insights into the VSIR result. First, genetically-predicted VSIR gene expression was associated with plasma levels of CXCL16 and CCL17, both of which are chemokines involved in immune responses^44,45^, and could mediate the association between *VSIR* and T1D. Second, *VSIR* was inversely associated with stool abundance of a strain of Blautia, and some have postulated a role for the gastrointestinal immune system in T1D pathogenesis^46^. Lower fecal levels of Blautia have been found in children with T1D^46^. Third, associations between higher genetically-predicted *VSIR* expression and basal cell carcinoma (BCC) may suggest possible safety concerns with VISTA agonism. This is likely due to loss of cancer immune surveillance, evidenced by increased incidence of BCC after immunosuppression initiation for renal transplantation^47^. A similar association for BCC exists for *CTLA4^48^*, a protein that also prevents T cell activation. Abatacept is a biologic drug that contains a CTLA-4 mimic currently approved for rheumatoid arthritis and has also been tested for T1D^49^. A meta-analysis of nine randomized trials did not detect a difference in BCC among those on Abatacept versus placebo^50^. Therefore, if Abatacept causes BCC, the effect is likely modest, which may also be the case for VISTA agonism. Nevertheless, our findings suggest the need for heightened surveillance of skin cancers if long-term VISTA agonism for T1D is attempted.

We also identified *P2RY12* as a novel therapeutic target to prevent or delay T1D. *P2RY12* is targeted by several approved drugs, including clopidogrel, that prevent secondary events after acute myocardial infarction and ischemic stroke. While this may not seem like a biologically relevant pathway for the pathogenesis of T1D, the result EBV protein EBNA-1 is intriguing, as EBV has been linked to development of several autoimmune diseases^51^, including T1D^52^. Notably, antibodies to EBV, and particularly EBNA-1, were found in higher percentage of newly diagnosed T1D compared to healthy controls^53^. Antibodies to ENBA1 cross-react with host Hepatic and Glial Cell Adhesion Molecule (GlialCAM) proteins, which may be a driver of the autoimmunity in multiple sclerosis^54^. Regarding existing functional work on *P2RY12*, our results corroborate studies that found *P2RY12* knockout mice are protected from autoimmune hepatitis^55^, and, importantly, had improved blood glucose and less damage to the islets of Langerhans in mice with induced T1D^56^. Further investigation into the role of *P2RY12* in the connection between EBV and T1D and a possible opportunity for use of *P2RY12* inhibition is warranted.

Several of our prioritized genes for T1D also influence risk of other autoimmune diseases, like *SESN3* and *CLNK*. Sestrin 3 (encoded by *SESN3*) is a stress-inducible protein that protects cells from oxidative damage. An analysis using human T cells found that the signal for rheumatoid arthritis (the same signal as one for T1D) in the *SESN3* region is driven by chromatin interactions that regulate *SESN3* expression^57^. Interestingly, our analysis suggested that *SESN3* may reduce T cell expression of CD3, a co-receptor essential for T cell activation and the target of teplizumab (a monoclonal antibody approved to delay the onset of stage 3 type 1 diabetes)^4^, and CD28, a key activator of T cells. One mouse knockout study of *CLNK*, a protein involved in immune signaling pathways, suggested an inhibitory function in natural killer cells^58^.

The major strength of this analysis was the use of eQTLs derived in isolated immune cells, which play an important role in the pathogenesis of T1D. Another strength is our use of conditional colocalization between eQTLs and both T1D and PheWAS outcomes to evaluate for confounding by LD. Lastly, we conducted follow-up analyses to assess for novelty, horizontal pleiotropy, and potential on-target effects or mediating phenotypes. There are also limitations to our study worth mentioning. First, gene expression may not directly correspond to levels of functional protein, and proteins, not genes, are the target of most therapeutics. For example, gene expression may be upregulated as a compensatory mechanism for a protein that lacks full function, which can falsely suggest that higher genetically-predicted gene expression increases disease risk, when in reality lower levels of a functional protein are instead associated with higher risk of disease. Second, gene expression may have a non-linear effect on T1D, and this may not be captured by summary-level MR, which assumes a linear relationship between exposure and outcome. Last, our analysis was restricted to those primarily of European ancestry.

In conclusion, we identified several novel genes with a potential causal relationship with T1D that could serve as targets for therapeutics to prevent or delay onset of T1D. Two such examples are *VSIR* and *P2RY12*, both of which have existing functional work demonstrating their role in autoimmune disease. We add supportive evidence using human genetic data, which is an important predictor in the success of a therapeutic reaching the market. Further work to investigate modulation of *VSIR*, *P2RY12,* and the other prioritized genes to prevent or delay T1D is warranted.

## Data Availability

All data produced in the present work are contained in the manuscript.

